# Incorporating Social Determinants of Health in the Prediction of Chronic Kidney Disease Progression in a National Cohort of US Veterans

**DOI:** 10.1101/2023.07.14.23292677

**Authors:** Karandeep Singh, Jennifer Bragg-Gresham, Yun Han, Brenda Gillespie, William Weitzel, Susan Crowley, Rajiv Saran

## Abstract

**Background:** As new therapies such as Sodium-Glucose Cotransporter-2 (SGLT2) inhibitors become available that may be effective earlier in the course of chronic kidney disease (CKD), newer risk prediction tools are required earlier in the course of CKD, and also to evaluate whether social determinants of health play a role in addition to individual-level risk factors.

**Methods:** We examined CKD progression among US Veterans who had known CKD in the VA health system using data from 2006-2016. CKD progression was defined based on two separate outcomes: 1) rapid CKD progression based on the eGFR slope < −3.7 mL/min/1.73m^2^ as a binary outcome, and 2) time-to-end stage kidney disease (ESKD) as a survival outcome. Veterans whose eGFR values were declining more steeply than −3.7 per year were considered “fast progressors,” representing 9.8% of the overall cohort. ESKD was identified by linking the VA data with US Renal Data System (USRDS) data, a national ESKD registry. After randomly dividing the dataset into a training, tuning, and testing set, tree ensemble models were trained and evaluated.

**Results:** We identified 1,550,526 patients meeting inclusion criteria, of which 930,615 patients were assigned to a training cohort, 309,831 to a tuning cohort, and 310,044 to a testing cohort. Tree ensemble models predicted fast progression with a C-statistic of 0.79 and time to ESKD with a C-statistic of 0.90. Baseline eGFR was the most important variable in predicting both outcomes, though social determinants constituted more of the important variables in rapid progression

**Conclusions:** CKD progression can be accurately predicted, though the predictors differ for fast progression and ESKD onset.

## INTRODUCTION

The primary therapeutic goals in patients with known chronic kidney disease (CKD) are to prevent kidney disease progression, manage comorbidities and complications such as cardiovascular disease, and in later stages, to prepare patients to safely transition to renal replacement therapies. While nephrologists are well-suited to identify and manage higher-risk CKD patients, their risk estimates for individual patients are often miscalibrated. For example, in a study of patients with chronic kidney disease (CKD) stage 3-5, nephrologists overestimated patients’ two-year risk of progressing to end stage kidney disease (ESKD):^1^ at predicted risk levels of 20% and 60%, the observed risks of progression were only 10% and 30%, respectively. Kidney Disease Improving Global Outcomes (KDIGO) recommends that patients be referred to nephrology when their risk of ESKD exceeds 10-20% and points to existing tools, such as the Kidney Failure Risk Equation (KFRE),^2^ to support this recommendation.

The KFRE is a Cox regression model that predicts the time-to-ESKD in patients with known CKD. It has been validated and recalibrated in a number of international settings, and it has been shown to outperform nephrologists in estimating patients’ risk.^1^ Because it was trained with dialysis and kidney transplant as target outcomes, it is most useful in identifying patients requiring preparation for renal replacement therapy. However, as new therapies such as Sodium-Glucose Cotransporter-2 (SGLT2) inhibitors, become available and be effective earlier in the course of CKD, new tools are required that are able to differentiate risk sooner. The slope of estimated glomerular filtration rate (eGFR) has previously been demonstrated to be a useful clinical marker of progression and has the advantage of being calculable in earlier CKD stages. Predicting the eGFR slope may be more clinically relevant in early CKD,^3, 4^ but the rate of early CKD progression may also be driven by social determinants of health,^5–7^ data for which are often not available in the electronic health record.

In this study, we sought to evaluate whether CKD progression can be accurately predicted when social determinants of health are incorporated into prediction models. While social determinants were not directly measured, we used geospatial data in a national cohort of US Veterans to link patient-level data to census-tract-level social determinants of health information derived from publicly available data sources. We evaluated the ability of models to predict fast progression (as determined by eGFR slope) and time to ESKD.

## METHODS

### Data source and study population

Our study used data from a national VA cohort drawing on data from 118 VA hospitals and their associated outpatient practices. We have complied with all relevant ethical regulations. The study was approved by the Institutional Review Boards of the VA Ann Arbor Healthcare System and the University of Michigan Medical School, and the need for informed consent was waived.

We examined CKD progression among US Veterans who had known CKD in the VA health system. Data were obtained from 2006-2016, and the presence of CKD was defined based on any of the following: any measure of eGFR value < 60 mL/min/1.73m^2^ using the CKD-EPI formula,^8^ a urine albumin/creatinine ratio (ACR) >30mg/g of creatinine, or a CKD diagnosis by ICD-9 or ICD-10 codes. Patients were excluded if they had fewer than five eGFR values available over the period.

### Outcomes

CKD progression was defined based on two separate outcomes: 1) rapid CKD progression based on the eGFR slope < −3.7 mL/min/1.73m^2^ as a binary outcome, and 2) time-to-ESKD as a survival outcome. Veterans whose eGFR values were declining more steeply than −3.7 per year were considered “fast progressors” because this represented a slope that was 1 standard deviation more extreme than the mean eGFR slope. Of the overall cohort, 9.8% were considered fast progressors by this definition. The outcome of ESKD was identified by analysis of the linked VA data with the national ESKD Registry data, i.e., the US Renal Data System (USRDS).

### Predictors

For each patient, patient-level variables were collected from the VA database along with social determinants of health obtained from public data sources linked at the census-tract and county level. Patient-level variables included demographic variables (age, sex, race, ethnicity), comorbidities, social history (smoking, alcohol use, and drug use), systolic and diastolic blood pressure values, laboratory data (serum creatinine based - baseline eGFR, baseline urine ACR, blood urea nitrogen, hematocrit, hemoglobin A1c, glucose), and exposure to potential and known nephrotoxins (including antibiotics, ACE inhibitors, antiretrovirals, non-steroidal anti-inflammatory medications, anti-angiogenesis medications, proton pump inhibitors, and lithium). Race and ethnicity were primarily included to understand their role in predicting CKD progression, and their relationship to social determinants of health. Combined with the social determinants of health predictors, the models considered 192 predictor variables.

### Social determinants of health

Social determinants may play a role in the rate of CKD progression by affecting access to care, quality of care, and overall health. To examine their role, we included multiple variables related to social determinants of health. These included variables collected at the patient level relating to access to care (e.g., drive time and drive distance to the nearest primary, secondary, and tertiary care VA facilities), healthcare utilization (e.g., number of laboratory tests performed), and variables collected at either the census-tract or county levels. These were linked to public data sources by geocoding patient addresses to census tract and county levels (depending on the data source). Linked variables included neighborhood affluence, density of grocery stores and fast foods, whether a patient’s neighborhood is considered a food desert, civic service density, environmental pollution (median PM_2.5_ levels), and the percentage of nearby buildings constructed in each decade.

### Model derivation and validation

We used separate tree ensemble models to predict the two outcomes: fast eGFR progression and time-to-ESKD. Gradient-boosted decision trees (GBDTs) were used for fast progression, and random survival forests were used for time-to-ESKD (details in Supplementary Methods).

For each of the models, patient data were randomly divided into training, tuning, and test cohorts. The tuning cohort was used to decide on early stopping for the GBDT models. For both models, tree ensemble models were trained on the training data and evaluated in the testing data.

### Variable importance

For the GBDT model, variable importance was calculated for each variable using equation 45 from Friedman 2001 as implemented in the *h2o* R package.^9^ For the random survival forest model, variable importance was calculated using the sum of test statistics impurity measure as implemented in the *ranger* R package.^10^ Relationships between individual variables and outcomes were explored using partial dependence plots and Shapley summary plots.

### Handling of missing data

Missing values were mean-imputed, and dummy variables were created to indicate missingness in the original data. This is because while the *h2o* R package tree ensemble implementation supports missing values, the *ranger* R package random survival forest implementation does not.

## RESULTS

Among patients with known CKD, 3,237,113 patients were identified who had available laboratory data to calculate eGFR slope. After excluding extreme values of slope (likely due to lab errors) and restricting to patients with more than 5 labs available to calculate eGFR slope over a maximum follow-up period of 10 years, 1,550,526 patients remained, of which 930,615 patients were randomly assigned to a training cohort, 309,831 to a tuning cohort, and 310,044 to a testing cohort. Baseline characteristics of the patient population, stratified by rapid progression, are provided in **Table 1** and **Supplementary Table 1**.

**Table 1.** Comparisons of Means/Percentages of Characteristics in Slow vs. Fast Progressors.

| Variable | Slow Progressors | Fast Progressors | p-value |
| --- | --- | --- | --- |
| ***Demographics*** |  |  |  |
| Age (years) | 66.6 | 62.8 | <0.0001 |
| Male | 95.8% | 95.9% | 0.0049 |
| White Race | 60.7% | 58.4% | <0.0001 |
| Black race | 11.3% | 19.8% | <0.0001 |
| Asian Race | 0.32% | 0.54% | <0.0001 |
| American Indian/AK Native | 0.37% | 0.60% | <0.0001 |
| Pacific Islander/HI Native | 0.55% | 0.64% | <0.0002 |
| Race unknown | 22.8% | 14.0% | <0.0001 |
| Hispanic ethnicity | 4.0% | 6.0% | <0.0001 |
| ***Comorbidities*** |  |  |  |
| Anemia | 55.7% | 60.5% | <0.0001 |
| Cardiovascular Disease | 17.6% | 20.9% | <0.0001 |
| Diabetes | 35.8% | 63.8% | <0.0001 |
| Hypertension | 49.9% | 59.2% | <0.0001 |

### Rapid eGFR Progression

The tree ensemble model had a C-statistic of 0.79 in the testing cohort in predicting rapid eGFR progression. The most important 20 variables are shown in Table 2.

**Table 2.**
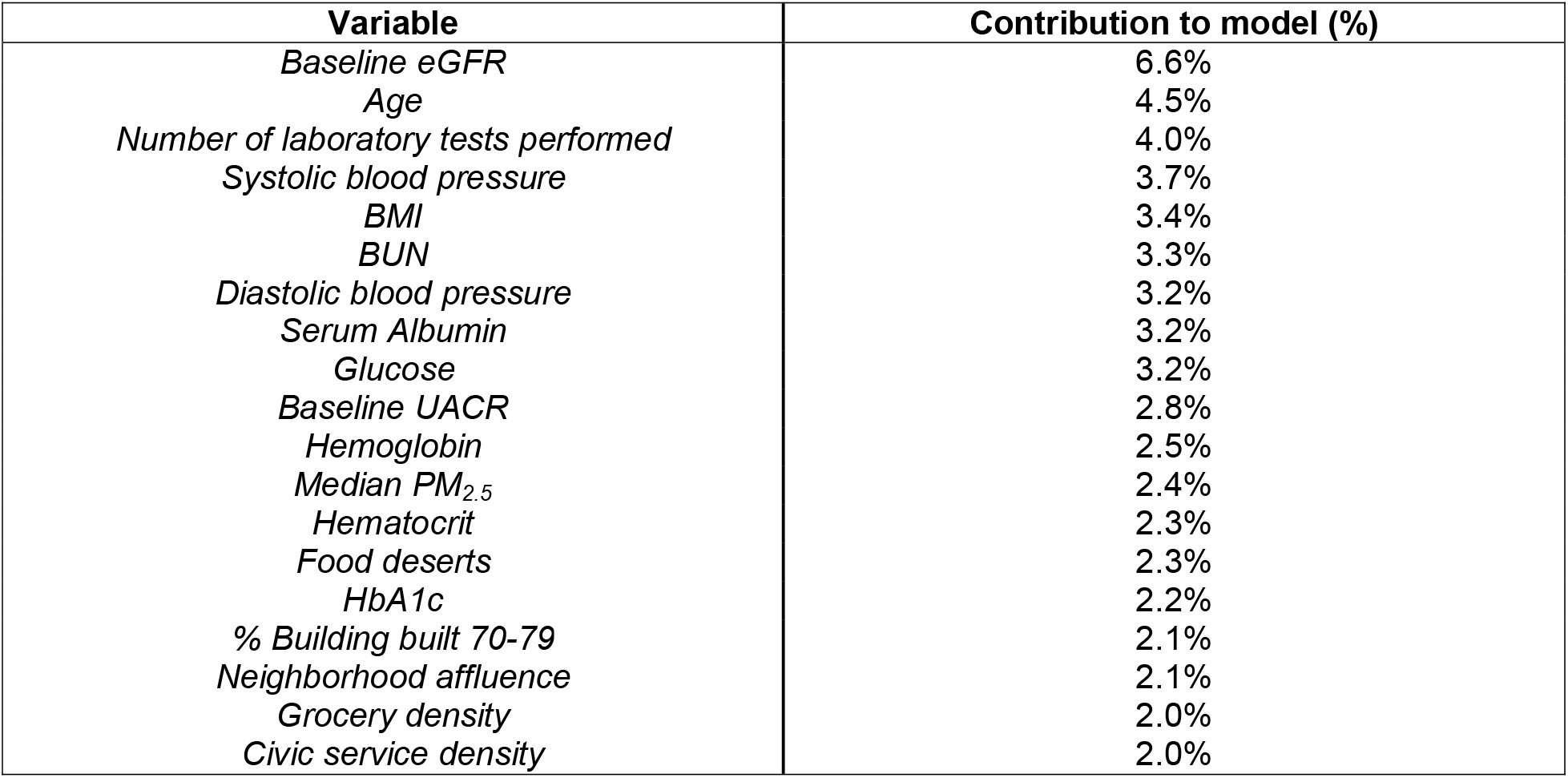
Variable importance for predicting “fast progressors” among patients with known CKD.

| **Variable** | **Contribution to model (%)** |
| --- | --- |
| *Baseline eGFR* | 6.6% |
| *Age* | 4.5% |
| *Number of laboratory tests performed* | 4.0% |
| *Systolic blood pressure* | 3.7% |
| *BMI* | 3.4% |
| *BUN* | 3.3% |
| *Diastolic blood pressure* | 3.2% |
| *Serum Albumin* | 3.2% |
| *Glucose* | 3.2% |
| *Baseline UACR* | 2.8% |
| *Hemoglobin* | 2.5% |
| *Median PM<sub>2.5</sub>* | 2.4% |
| *Hematocrit* | 2.3% |
| *Food deserts* | 2.3% |
| *HbA1c* | 2.2% |
| *% Building built 70-79* | 2.1% |
| *Neighborhood affluence* | 2.1% |
| *Grocery density* | 2.0% |
| *Civic service density* | 2.0% |

Baseline eGFR was the most important contributor to the model identifying fast progressors, followed by age and the number of laboratory tests performed. Among the social determinants of health, the most important predictors were median PM_2.5_ (a measure of environmental pollution), whether the patient’s neighborhood was a food desert, the percentage of nearby buildings built in the 1970s, neighborhood affluence, the density of grocery stores, and civic service density.

Patients’ risk of rapid progression increases as their baseline eGFR increases based on a partial dependence plot (**Figure 1**). This finding is similar in the Shapley summary plot (**Figure 2**), which shows that the highest risk of rapid progression (towards the right side of the plot) generally occurs when the baseline eGFR values are high (depicted as red dots).

**Figure 1.**
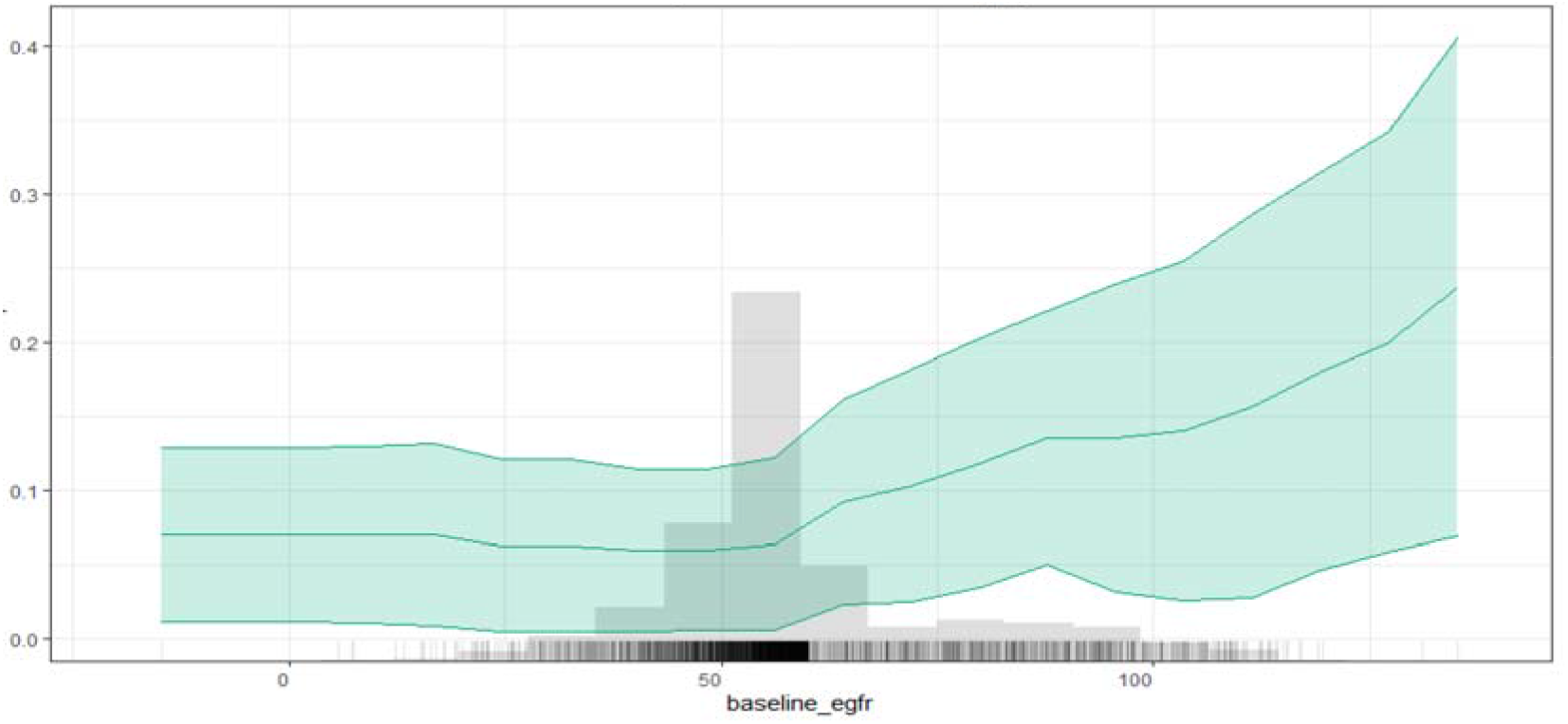
Partial dependence plot showing relationship between baseline eGFR and rapid CKD progression.

**Figure 2.**
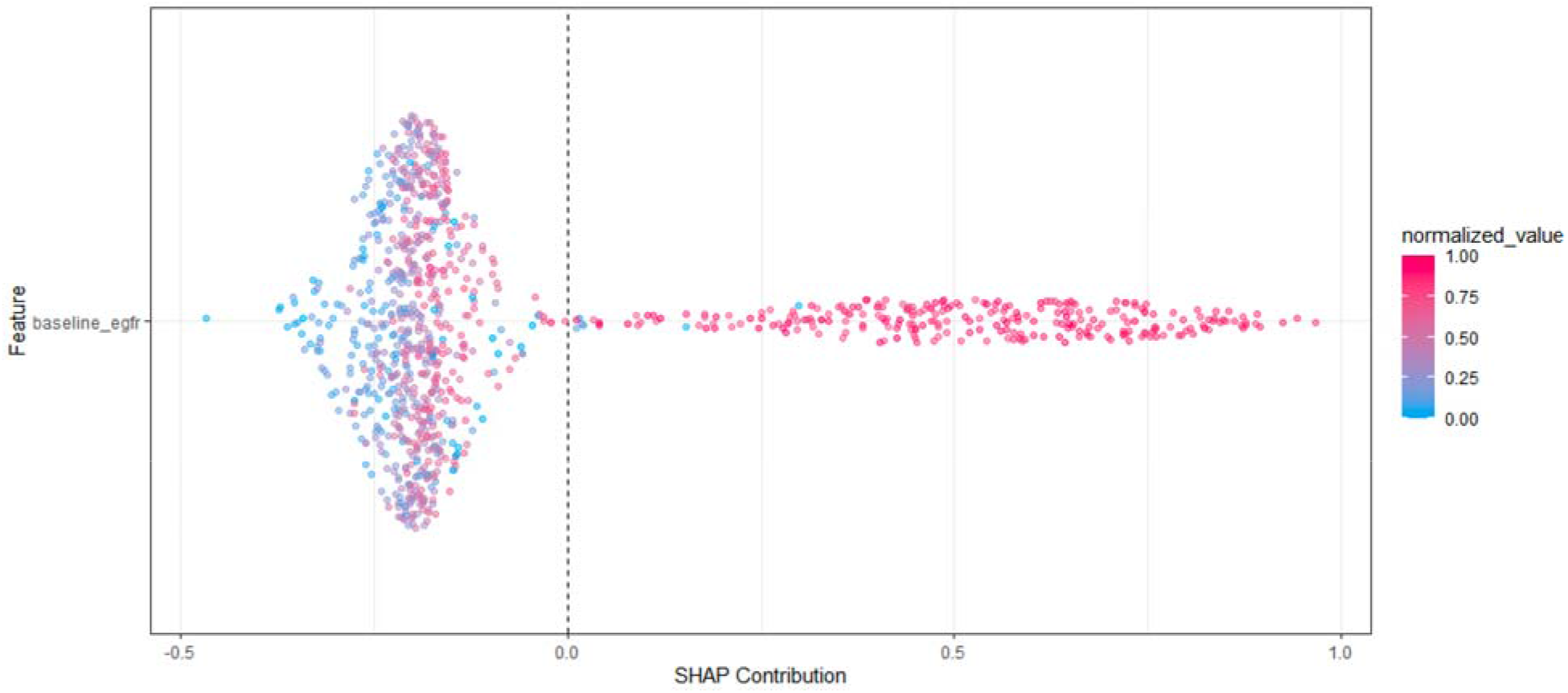
Shapley summary plot dependence plot showing relationship between baseline eGFR and rapid CKD progression.

### Predicting time-to-ESKD

The tree ensemble model had a C-statistic of 0.90 in the testing cohort in predicting time to ESRD. The most important 20 variables are shown in Table 3.

**Table 3.** Variable importance for predicting time-to-ESKD among patients with known CKD.

| Variable | Relative importance (out of 100%) |
| --- | --- |
| *Baseline eGFR* | 11% |
| *Outpatient blood urea nitrogen lab* | 7.9% |
| *Nephrology visit (binary)* | 4.8% |
| *Diagnosis of chronic kidney disease* | 3.3% |
| *Baseline urinary albumin/creatinine ratio* | 2.5% |
| *Outpatient hematology lab* | 2.2% |
| *Outpatient blood urea nitrogen (different source)* | 2.1% |
| *Outpatient hematocrit lab* | 1.9% |
| *Systolic blood pressure (mean)* | 1.8% |
| *Age* | 1.7% |
| *Systolic blood pressure in the prior year* | 1.7% |
| *Systolic blood pressure 2 years ago* | 1.7% |
| *Race* | 1.5% |
| *Outpatient hemoglobin A1c* | 1.4% |
| *Outpatient glucose* | 1.4% |
| *Diastolic blood pressure 2 years ago* | 1.1% |
| *Diastolic blood pressure in the past year* | 1.1% |
| *Diastolic blood pressure (mean)* | 1.1% |
| *Body mass index* | 1.0% |
| *Any evidence of CKD (narrow definition)* | 0.98% |

The most important variable in predicting time-to-ESKD was baseline eGFR, followed by the most recent outpatient blood urea nitrogen level and whether the patient had previously seen a nephrologist. The only social determinant of health among the top 20 variables was the presence of a nephrology visit, suggesting that social determinants might play a lesser role in predicting the time to ESKD, although this cannot be stated with certainty.

## DISCUSSION

In this study of US Veterans using clinical factors and social determinants of health in the prediction of CKD progression, we found that tree ensemble models can accurately predict the progression of CKD, with C-statistics of 0.79 for fast progression (earlier in the CKD continuum) and 0.90 for time to ESKD (later in the course of CKD). While baseline eGFR is the most important variable in predicting both outcomes, its relationship to the two outcomes is different. While a lower baseline eGFR indicates a higher risk of progression of ESKD, we found that it appears ‘protective’ against a rapid decline in eGFR based on slope. Although counterintuitive, we speculate that this might be related to high values of eGFR (>100) represent glomerular hyperfiltration which is a far less pronounced in advanced stages of CKD. When early values of eGFR are high, future values of eGFR will commonly be lower both due to the fact that glomerular hyperfiltration often predicts subsequent kidney damage with progressive decline in eGFR.

Beyond traditional risk factors for CKD progression, we also found social determinants of health to be important contributors to predicting risk based on tree ensemble variable importance. Important social determinants in predicting fast progressors included the number of laboratory tests performed, median PM_2.5_, food deserts, the percentage of nearby buildings built in the 1970s, neighborhood affluence, grocery density, and civic service density. Social determinants were less relevant to the prediction of EKSD onset, where only the presence of a nephrology visit was considered in the top 20 important variables. This finding is interesting because it suggests that social determinants are more relevant earlier in the course of CKD, which is where eGFR slope is more relevant. This may be because earlier eGFR decline is more modifiable, and changing the diet (i.e., healthy food) may have more of an impact in early CKD. The number of laboratory tests may be indicative of the clinical need to monitor kidney function more frequently as well as greater eGFR variability and may not necessarily be reflective only of social factors. However, the ability to undergo multiple laboratory tests is suggestive of having both transportation to a laboratory testing facility and sufficient healthcare coverage to afford multiple tests.

Our observation that social factors were less relevant to predicting ESKD onset is not surprising. Even in the kidney failure risk equation (KFRE), a 4-variable model achieved a C-statistic of 0.91,^2^ suggesting that demographic and biological risk factors capture a substantial degree of the risk of ESKD onset. Our model, which considered a much larger number of predictors, achieved a similar C-statistic of 0.90, suggesting that social determinants may play less of a role, although their role in ensuring progression in earlier stages of the disease is not excluded, and may be confounded by the advancing nature of the disease in later stages. The patients identified as high risk by the KFRE are generally those with lower eGFRs and thus are closer to the endpoint of renal replacement therapy. Thus, it may be too late for high-risk patients, i.e., those closer to the endpoint of renal replacement therapy, for social determinants to substantially appear to affect the risk, as they have already caused the most damage in earlier stages of the disease. Of the social factors we identified, having seen a nephrologist was the most important one. This is because the decision to start renal replacement therapy is typically made by a nephrologist and thus may represent an indicator that CKD has been clinically recognized, and that the patient has sufficient healthcare coverage to visit a nephrologist. This may also represent a case of ‘confounding by indication’ for ESKD treatment.

Our study has limitations. While our study was conducted in a large national cohort of US Veterans, factors that affect access to care in Veterans may be different from those affecting other populations, especially those relating to healthcare coverage. For example, the barriers to see a nephrologist may be lower or different in VA care settings as compared to other care settings, which could have affected our findings. While social determinants of health are generally well-captured by census tracts, there may be heterogeneity among social factors even within census tracts, which we did not capture as we did not measure social determinants at the patient level.

Despite these limitations, our finding of differing risk factors of fast progression and ESKD onset suggest that interventions to consider in these groups may be different. Because social factors are more important in fast progression—an outcome that occurs earlier in the course of CKD—interventions on social factors may potentially modify risk in early CKD progression.

## Data Availability

All data produced in the present work are contained in the manuscript

## SUPPLEMENTARY MATERIAL

### Supplementary Methods

#### Selection of tree ensemble models

Tree ensemble models were selected because they have been shown to perform similarly to or modestly better than regression models for kidney-related outcomes. There are multiple implementations of tree ensemble models in R. Tree ensemble models are non-parametric, include interactions, and are computationally efficient. We selected gradient-boosted decision trees (GBDTs) to predict rapid progression (as determined using a slope-based threshold) because GBDTs are more computationally efficient than random forests in their ability to achieve similar performance using fewer splits. For this, we selected the *h2o* R package implementation of GBDTs due to its ability to use multicore parallel processing while constraining the computer processing unit (CPU) usage, which was a key need when performing analyses on the VA’s VINCI platform. For the time-to-ESKD outcome, existing GBDT implementations did not support survival outcomes in R, so we opted for random survival forests (as implemented in the *ranger* R package).

A random forest survival model is similar to a Cox model in that it can model survival outcomes in the presence of censoring. A random forest consists of a set of independently fit decision trees. Each decision tree is fit on a bootstrap (or subsample) of the original training data. Each tree consists of a series of nodes, where a binary split is chosen for a variable after evaluating a number of candidate splits after considering a random subset of variables to split upon. This process is repeated recursively for each child node until either no further splitting is possible or a prespecified maximum depth is reached. Each terminal node in the decision tree is known as a “leaf node” and contains the predicted value of the outcome for the node path, which defines the path from the root node (first split) to the leaf node. Different patients are assigned to different node paths based on their underlying values for the predictors, and thus each tree generates a different prediction for each patient. These tree-level predictors are averaged across the entire forest to obtain a random forest prediction.

**Supplementary Table 1.**
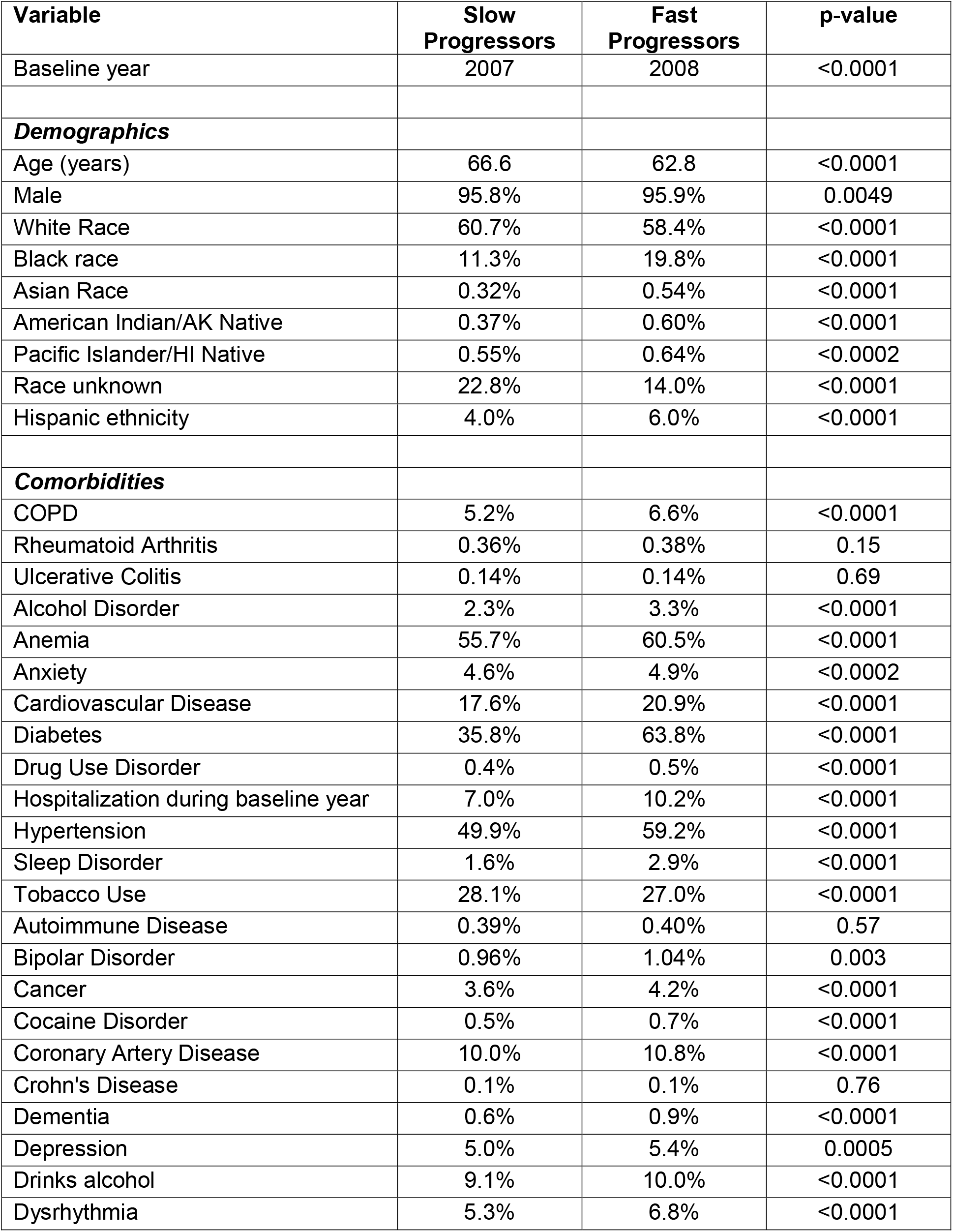
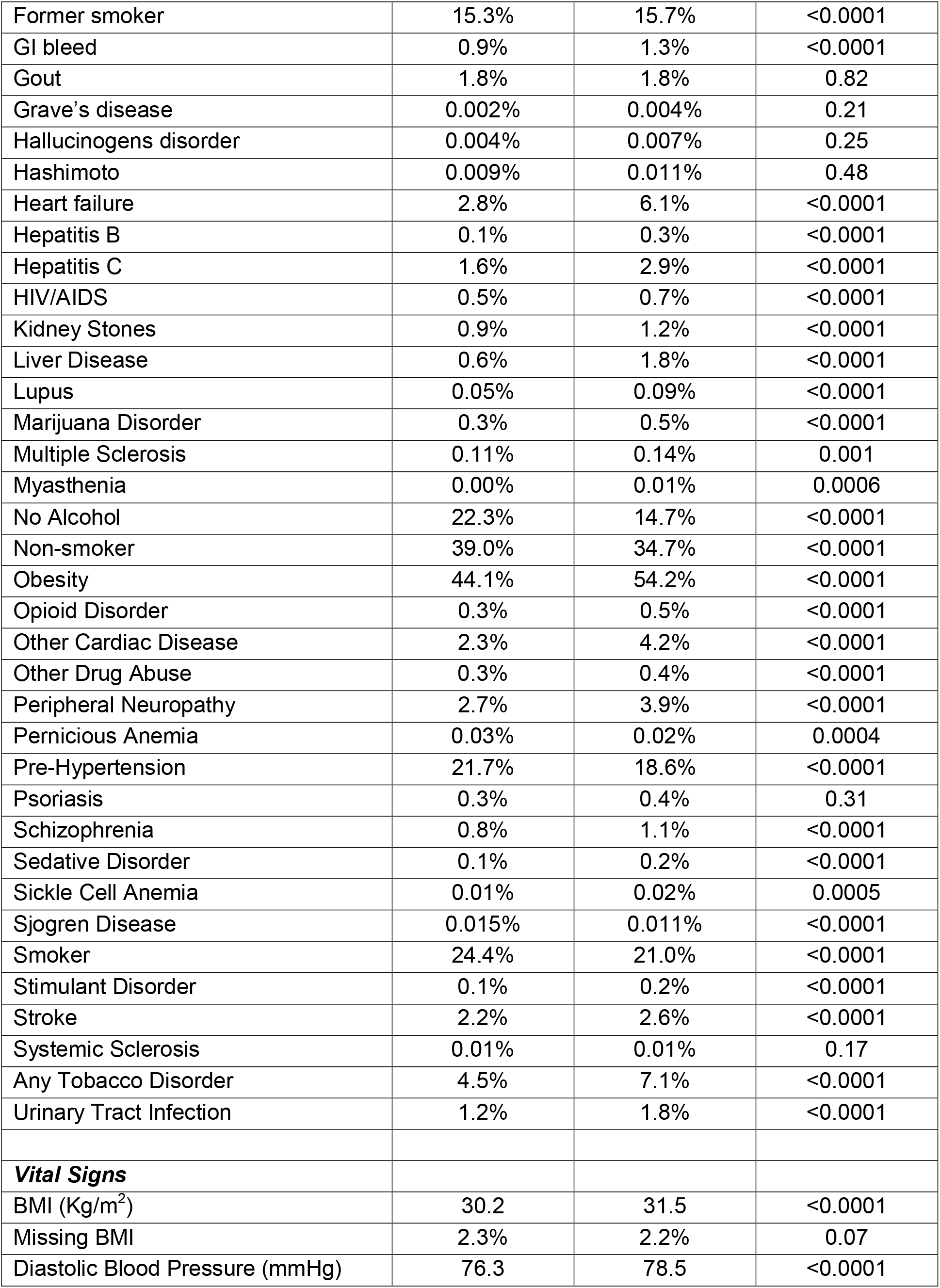
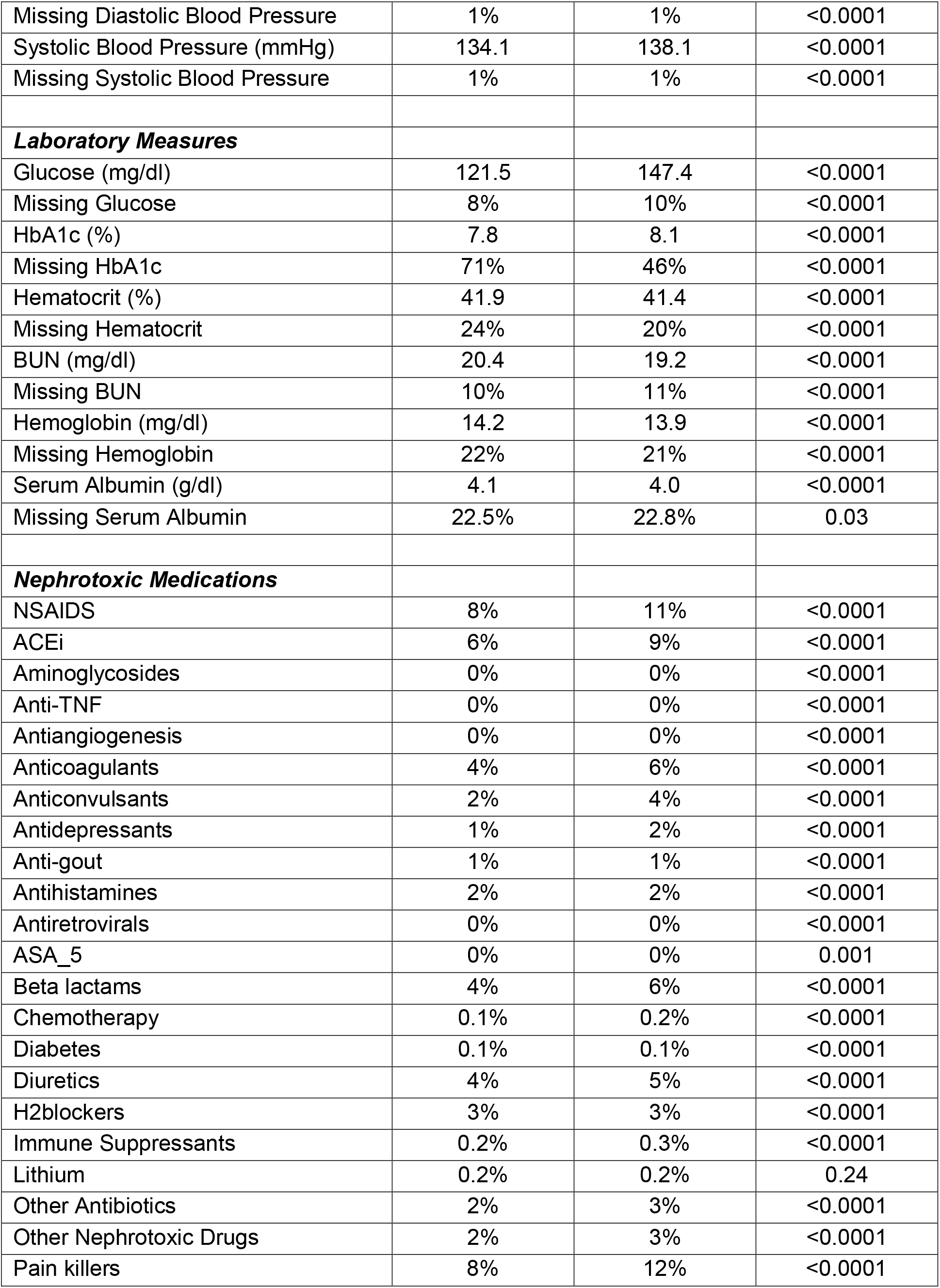
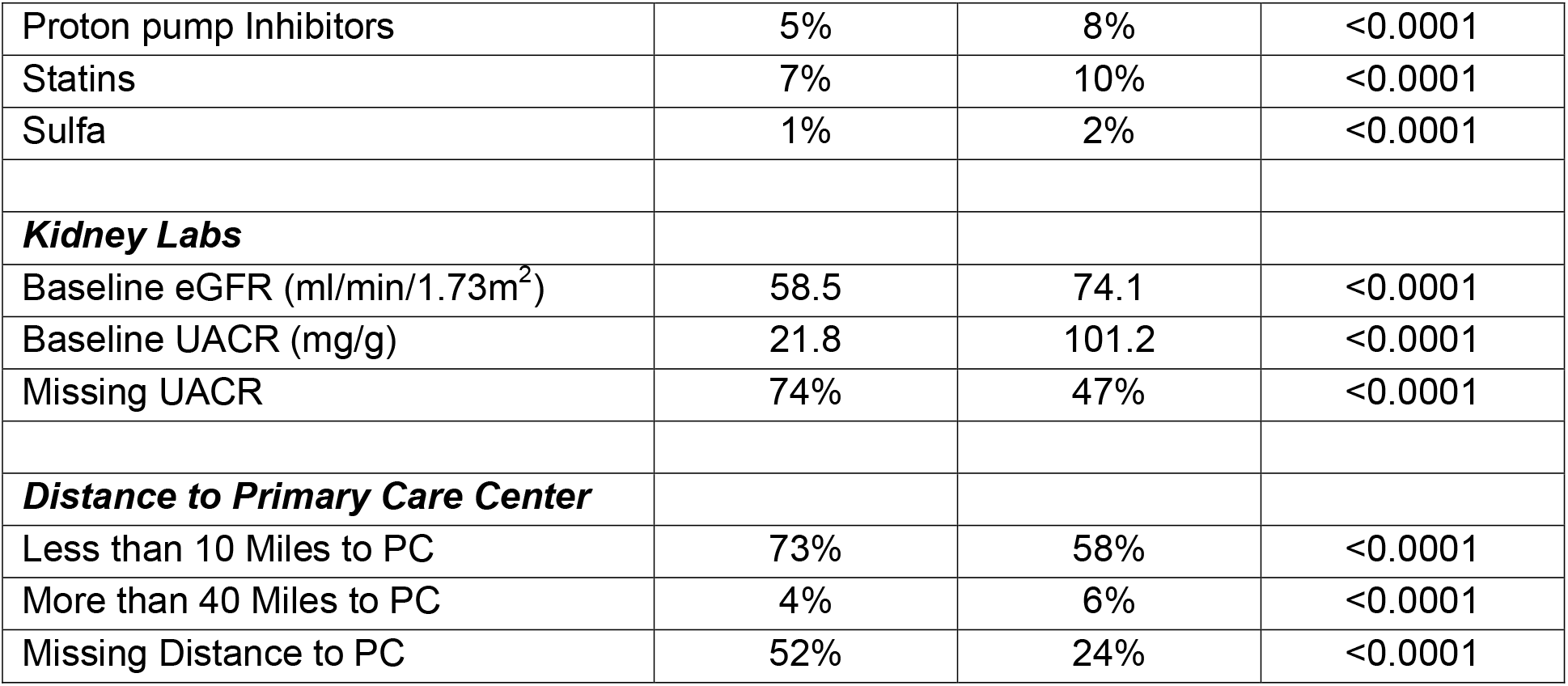
Comparisons of Means/Percentages of Characteristics in Slow vs. Fast Progressors. A separate comparison is made for each variable.

| **Variable** | **Slow Progressors** | **Fast Progressors** | **p-value** |
| --- | --- | --- | --- |
| Baseline year | 2007 | 2008 | <0.0001 |
| ***Demographics*** |  |  |  |
| Age (years) | 66.6 | 62.8 | <0.0001 |
| Male | 95.8% | 95.9% | 0.0049 |
| White Race | 60.7% | 58.4% | <0.0001 |
| Black race | 11.3% | 19.8% | <0.0001 |
| Asian Race | 0.32% | 0.54% | <0.0001 |
| American Indian/AK Native | 0.37% | 0.60% | <0.0001 |
| Pacific Islander/HI Native | 0.55% | 0.64% | <0.0002 |
| Race unknown | 22.8% | 14.0% | <0.0001 |
| Hispanic ethnicity | 4.0% | 6.0% | <0.0001 |
| ***Comorbidities*** |  |  |  |
| COPD | 5.2% | 6.6% | <0.0001 |
| Rheumatoid Arthritis | 0.36% | 0.38% | 0.15 |
| Ulcerative Colitis | 0.14% | 0.14% | 0.69 |
| Alcohol Disorder | 2.3% | 3.3% | <0.0001 |
| Anemia | 55.7% | 60.5% | <0.0001 |
| Anxiety | 4.6% | 4.9% | <0.0002 |
| Cardiovascular Disease | 17.6% | 20.9% | <0.0001 |
| Diabetes | 35.8% | 63.8% | <0.0001 |
| Drug Use Disorder | 0.4% | 0.5% | <0.0001 |
| Hospitalization during baseline year | 7.0% | 10.2% | <0.0001 |
| Hypertension | 49.9% | 59.2% | <0.0001 |
| Sleep Disorder | 1.6% | 2.9% | <0.0001 |
| Tobacco Use | 28.1% | 27.0% | <0.0001 |
| Autoimmune Disease | 0.39% | 0.40% | 0.57 |
| Bipolar Disorder | 0.96% | 1.04% | 0.003 |
| Cancer | 3.6% | 4.2% | <0.0001 |
| Cocaine Disorder | 0.5% | 0.7% | <0.0001 |
| Coronary Artery Disease | 10.0% | 10.8% | <0.0001 |
| Crohn's Disease | 0.1% | 0.1% | 0.76 |
| Dementia | 0.6% | 0.9% | <0.0001 |
| Depression | 5.0% | 5.4% | 0.0005 |
| Drinks alcohol | 9.1% | 10.0% | <0.0001 |
| Dysrhythmia | 5.3% | 6.8% | <0.0001 |
| Former smoker | 15.3% | 15.7% | <0.0001 |
| GI bleed | 0.9% | 1.3% | <0.0001 |
| Gout | 1.8% | 1.8% | 0.82 |
| Grave's disease | 0.002% | 0.004% | 0.21 |
| Hallucinogens disorder | 0.004% | 0.007% | 0.25 |
| Hashimoto | 0.009% | 0.011% | 0.48 |
| Heart failure | 2.8% | 6.1% | <0.0001 |
| Hepatitis B | 0.1% | 0.3% | <0.0001 |
| Hepatitis C | 1.6% | 2.9% | <0.0001 |
| HIV/AIDS | 0.5% | 0.7% | <0.0001 |
| Kidney Stones | 0.9% | 1.2% | <0.0001 |
| Liver Disease | 0.6% | 1.8% | <0.0001 |
| Lupus | 0.05% | 0.09% | <0.0001 |
| Marijuana Disorder | 0.3% | 0.5% | <0.0001 |
| Multiple Sclerosis | 0.11% | 0.14% | 0.001 |
| Myasthenia | 0.00% | 0.01% | 0.0006 |
| No Alcohol | 22.3% | 14.7% | <0.0001 |
| Non-smoker | 39.0% | 34.7% | <0.0001 |
| Obesity | 44.1% | 54.2% | <0.0001 |
| Opioid Disorder | 0.3% | 0.5% | <0.0001 |
| Other Cardiac Disease | 2.3% | 4.2% | <0.0001 |
| Other Drug Abuse | 0.3% | 0.4% | <0.0001 |
| Peripheral Neuropathy | 2.7% | 3.9% | <0.0001 |
| Pernicious Anemia | 0.03% | 0.02% | 0.0004 |
| Pre-Hypertension | 21.7% | 18.6% | <0.0001 |
| Psoriasis | 0.3% | 0.4% | 0.31 |
| Schizophrenia | 0.8% | 1.1% | <0.0001 |
| Sedative Disorder | 0.1% | 0.2% | <0.0001 |
| Sickle Cell Anemia | 0.01% | 0.02% | 0.0005 |
| Sjogren Disease | 0.015% | 0.011% | <0.0001 |
| Smoker | 24.4% | 21.0% | <0.0001 |
| Stimulant Disorder | 0.1% | 0.2% | <0.0001 |
| Stroke | 2.2% | 2.6% | <0.0001 |
| Systemic Sclerosis | 0.01% | 0.01% | 0.17 |
| Any Tobacco Disorder | 4.5% | 7.1% | <0.0001 |
| Urinary Tract Infection | 1.2% | 1.8% | <0.0001 |
| **Vital Signs** |  |  |  |
| BMI (Kg/m<sup>2</sup>) | 30.2 | 31.5 | <0.0001 |
| Missing BMI | 2.3% | 2.2% | 0.07 |
| Diastolic Blood Pressure (mmHg) | 76.3 | 78.5 | <0.0001 |
| Missing Diastolic Blood Pressure | 1% | 1% | <0.0001 |
| Systolic Blood Pressure (mmHg) | 134.1 | 138.1 | <0.0001 |
| Missing Systolic Blood Pressure | 1% | 1% | <0.0001 |
| **Laboratory Measures** |  |  |  |
| Glucose (mg/dl) | 121.5 | 147.4 | <0.0001 |
| Missing Glucose | 8% | 10% | <0.0001 |
| HbA1c (%) | 7.8 | 8.1 | <0.0001 |
| Missing HbA1c | 71% | 46% | <0.0001 |
| Hematocrit (%) | 41.9 | 41.4 | <0.0001 |
| Missing Hematocrit | 24% | 20% | <0.0001 |
| BUN (mg/dl) | 20.4 | 19.2 | <0.0001 |
| Missing BUN | 10% | 11% | <0.0001 |
| Hemoglobin (mg/dl) | 14.2 | 13.9 | <0.0001 |
| Missing Hemoglobin | 22% | 21% | <0.0001 |
| Serum Albumin (g/dl) | 4.1 | 4.0 | <0.0001 |
| Missing Serum Albumin | 22.5% | 22.8% | 0.03 |
| **Nephrotoxic Medications** |  |  |  |
| NSAIDS | 8% | 11% | <0.0001 |
| ACEi | 6% | 9% | <0.0001 |
| Aminoglycosides | 0% | 0% | <0.0001 |
| Anti-TNF | 0% | 0% | <0.0001 |
| Antiangiogenesis | 0% | 0% | <0.0001 |
| Anticoagulants | 4% | 6% | <0.0001 |
| Anticonvulsants | 2% | 4% | <0.0001 |
| Antidepressants | 1% | 2% | <0.0001 |
| Anti-gout | 1% | 1% | <0.0001 |
| Antihistamines | 2% | 2% | <0.0001 |
| Antiretrovirals | 0% | 0% | <0.0001 |
| ASA_5 | 0% | 0% | 0.001 |
| Beta lactams | 4% | 6% | <0.0001 |
| Chemotherapy | 0.1% | 0.2% | <0.0001 |
| Diabetes | 0.1% | 0.1% | <0.0001 |
| Diuretics | 4% | 5% | <0.0001 |
| H2blockers | 3% | 3% | <0.0001 |
| Immune Suppressants | 0.2% | 0.3% | <0.0001 |
| Lithium | 0.2% | 0.2% | 0.24 |
| Other Antibiotics | 2% | 3% | <0.0001 |
| Other Nephrotoxic Drugs | 2% | 3% | <0.0001 |
| Pain killers | 8% | 12% | <0.0001 |
| Proton pump Inhibitors | 5% | 8% | <0.0001 |
| Statins | 7% | 10% | <0.0001 |
| Sulfa | 1% | 2% | <0.0001 |
| ***Kidney Labs*** |  |  |  |
| Baseline eGFR (ml/min/1.73m<sup>2</sup>) | 58.5 | 74.1 | <0.0001 |
| Baseline UACR (mg/g) | 21.8 | 101.2 | <0.0001 |
| Missing UACR | 74% | 47% | <0.0001 |
| ***Distance to Primary Care Center*** |  |  |  |
| Less than 10 Miles to PC | 73% | 58% | <0.0001 |
| More than 40 Miles to PC | 4% | 6% | <0.0001 |
| Missing Distance to PC | 52% | 24% | <0.0001 |

